# Digital phenotyping from wearables using AI characterizes psychiatric disorders and identifies genetic associations

**DOI:** 10.1101/2024.09.23.24314219

**Authors:** Jason Liu, Beatrice Borsari, Yunyang Li, Susanna Liu, Yuan Gao, Xin Xin, Shaoke Lou, Matthew Jensen, Diego Garrido-Martín, Terril Verplaetse, Garrett Ash, Jing Zhang, Matthew J. Girgenti, Walter Roberts, Mark Gerstein

## Abstract

Psychiatric disorders are complex and influenced by both genetic and environmental factors. However, studying the full spectrum of these disorders is hindered by practical limitations on measuring human behavior. This highlights the need for novel technologies that can measure behavioral changes at an intermediate level between diagnosis and genotype. Wearable devices are a promising tool in precision medicine, since they can record physiological measurements over time in response to environmental stimuli and do so at low cost and minimal invasiveness. Here we analyzed wearable and genetic data from a cohort of the Adolescent Brain Cognitive Development study. We generated >250 wearable-derived features and used them as intermediate phenotypes in an interpretable AI modeling framework to assign risk scores and classify adolescents with psychiatric disorders. Our model identifies key physiological processes and leverages their temporal patterns to achieve a higher performance than has been previously possible. To investigate how these physiological processes relate to the underlying genetic architecture of psychiatric disorders, we also utilized these intermediate phenotypes in univariate and multivariate GWAS. We identified a total of 29 significant genetic loci and 52 psychiatric-associated genes, including ELFN1 and ADORA3. These results show that wearable-derived continuous features enable a more precise representation of psychiatric disorders and exhibit greater detection power compared to categorical diagnostic labels. In summary, we demonstrate how consumer wearable technology can facilitate dimensional approaches in precision psychiatry and uncover etiological linkages between behavior and genetics.

## Introduction

Psychiatric disorders of childhood and adolescence currently affect 1 in 7 youths in the United States and globally^1,2^. Externalizing disorders such as attention-deficit/hyperactivity disorder, and internalizing disorders such as anxiety, are among the most prevalent and represent a wide spectrum of dysfunctional behavior patterns^3^. Treatment barriers are complex and multifaceted but major contributors include our limited understanding of psychiatric phenotypes and difficulty identifying youth individuals that experience these disorders.

Traditionally, psychiatric disorders have been conceptualized as categorical macrophenotypes, based on clinical manifestations of a disease which are defined according to the number and type of symptoms, and the presence of distress or impairment^4–6^. While this has practical benefits in terms of reliability and ease of diagnosis, it poses several challenges to the research of these disorders, and consequently to the development of treatments. In fact, psychiatric disorders are complex and often comorbid, and this high degree of heterogeneity is not always accurately translated into categorical diagnosis labels, which may be defined by arbitrary cut-offs. Instead, intermediate phenotypes (i.e., quantitative traits that are positioned between genotype and macrophenotype) may better capture the heterogeneity potentially missed by existing diagnostic categories^7–9^. Additionally, genetic penetrance is expected to be higher for these intermediate phenotypes compared to macrophenotypes, enabling improved dissection of the genetic architecture underlying psychiatric disorders^10^. Nevertheless, many genome-wide association studies (GWAS) aimed at identifying genetic variants or biomarkers for psychiatric disorders do not consider these intermediate phenotypes and instead rely on dichotomised (i.e., binary) traits. In fact, identifying intermediate phenotypes with clinical and biological relevance remains a challenge^11^.

Therefore, to improve our understanding of psychiatric disorders it is important that we identify intermediate phenotypes that not only offer a more comprehensive representation of an individual’s behavior with respect to their environment, but also relate well with existing clinical definitions and aid in diagnosis. Once identified, these intermediate phenotypes can then be also used to guide more comprehensive studies to identify genetic associations and biomarkers that may ultimately improve precision treatments.

To achieve this goal, it is important to leverage new emerging technologies that can quantitatively assess an individual’s behavioral patterns^12^. Wearable sensors such as smartwatches collect data that reflect physical and physiological processes (e.g., movement, pulse, metabolic intake), and can be used to infer higher-order behavioral events (e.g., sleep, exercise) and their temporal dynamics. Because of the documented relationship between such higher-order behavioral events and mental health, and given their low cost and minimal invasiveness, wearable devices have emerged as promising tools for mental health monitoring and psychiatric evaluation^13–15^.

Therefore, wearable sensors have the potential for capturing intermediate phenotypes relevant to behavior and psychiatric disorders, ultimately enabling improved GWAS. However, significant computational challenges remain in generating intermediate phenotypes from wearable-derived data that describe the full spectrum of a given psychiatric disorder. Moreover, further curation of these intermediate phenotypes is necessary to identify genetic associations that have clinical and biological relevance.

To address these limitations, we developed an AI modeling framework that flexibly leverages data from wearable devices to generate intermediate phenotypes in the form of static and dynamic digital features. We establish the validity of these digital features as intermediate phenotypes by classifying externalizing and internalizing disorders with an accuracy beyond baseline expectation, and even surpassing the performance of some other gold-standard intermediate phenotypes such as fMRI measurements^16–19^. Interpretability modules in our AI framework enable us to identify key temporal and physiological insights between clinical diagnosis and digital features, further supporting the validity of using these wearable-derived features as intermediate phenotypes. We further curate these intermediate phenotypes and employ them in GWAS models to identify genetic associations and biomarkers that capture the continuous spectrum of psychiatric disorders and behavioral patterns. Finally, we identify 29 significant loci, several of which overlap previously reported genetic variants associated with behavioral traits and mental illnesses and are proximal to genes with a documented role in neurodevelopmental and psychiatric disorders.

In sum, this work shows how wearable devices can advance our understanding of psychiatric disorders by establishing a more objective and dimensional approach that can ultimately lead to improved treatments in precision psychiatry.

## Results

### Leveraging the Adolescent Brain Cognitive Development cohort

To improve our understanding of psychiatric disorders, we leveraged and analyzed a dataset from a cohort of US adolescents recruited by the NIH Adolescent Brain Cognitive Development Consortium (ABCD) project, consisting of clinical, wearable, and genetic data (**Fig. 1**). The ABCD cohort consists of a total of 11,878 adolescents (5682 males and 6196 females), of age between nine and fourteen years and belonging to four different ethnicities. We identified nine categories of psychiatric phenotypes, which were established using a gold standard parent diagnostic semi-structured interview (Kiddie Schedule for Affective Disorders and Schizophrenia-5)^20^. The healthy controls represented adolescents who did not meet the criteria for any of those nine psychiatric disorders. We defined these clinical labels as the categorical macrophenotypes in the study (**Fig. 1A-B**). Our modeling framework also utilized data from cognitive tests (e.g., NIH Toolbox) and behavioral checklists (**Fig. 2A**).

**Figure 1.**
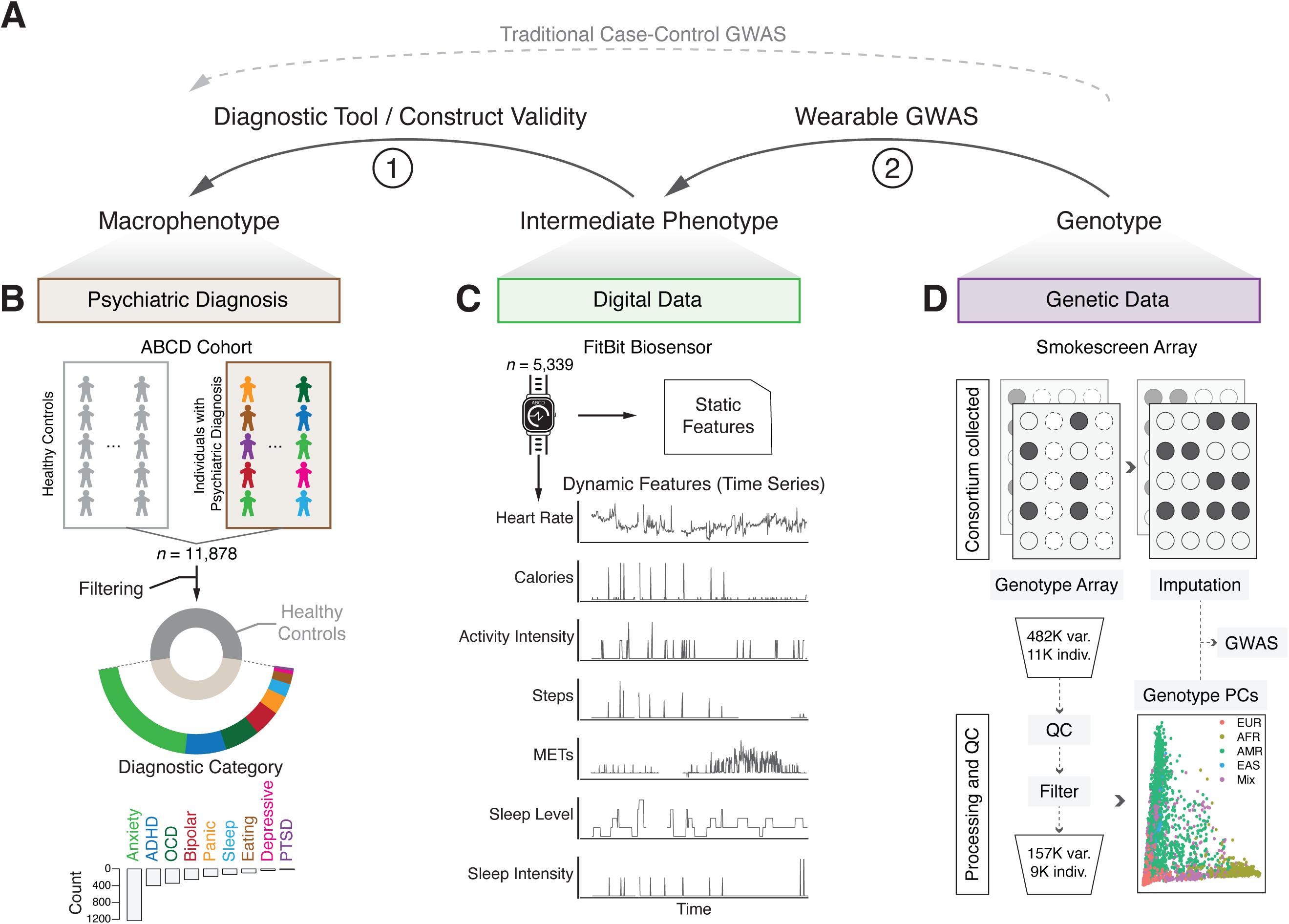
Leveraging clinical, digital, and genetic data of the ABCD cohort to improve characterization of psychiatric disorders. **A)** Framework schematic describing how intermediate phenotypes from wearable-derived data are leveraged to better understand the association between macrophenotype and genotype. The link between intermediate phenotype and macrophenotype serves as construct validity and aid in diagnostics. Wearable GWAS is performed through genotype-to-intermediate-phenotype association studies. **B)** The Adolescent Brain Cognitive Development (ABCD) cohort contains 11,878 individuals spanning nine different categorical macrophenotypes based on clinical diagnosis from the Kiddie Schedule for Affective Disorders and Schizophrenia-5. A breakdown of the counts of each disorder is shown in the bottom bar graph, with anxiety disorder and ADHD being the most prevalent. “Bipolar” refers to bipolar or psychotic disorders. **C)** Digital data from FitBit biosensors are collected for 5,339 individuals. The collected time series data are then processed into dynamic and static features, with information spanning various physiological and higher order processes. **D)** Genetic data are collected by the ABCD consortium through Smokescreen genotyping array. Imputed genotypes are used for downstream GWAS analyses. The genotype arrays are subjected to best-practice processing and QC to ensure included individuals and SNPs are of high quality. PCA performed on 8,791 individuals and 157,556 genotyped SNPs reveals distinct ancestral clusters across the cohort and the inferred genotype principal components (PCs) are used as covariates in downstream analyses.

**Figure 2.**
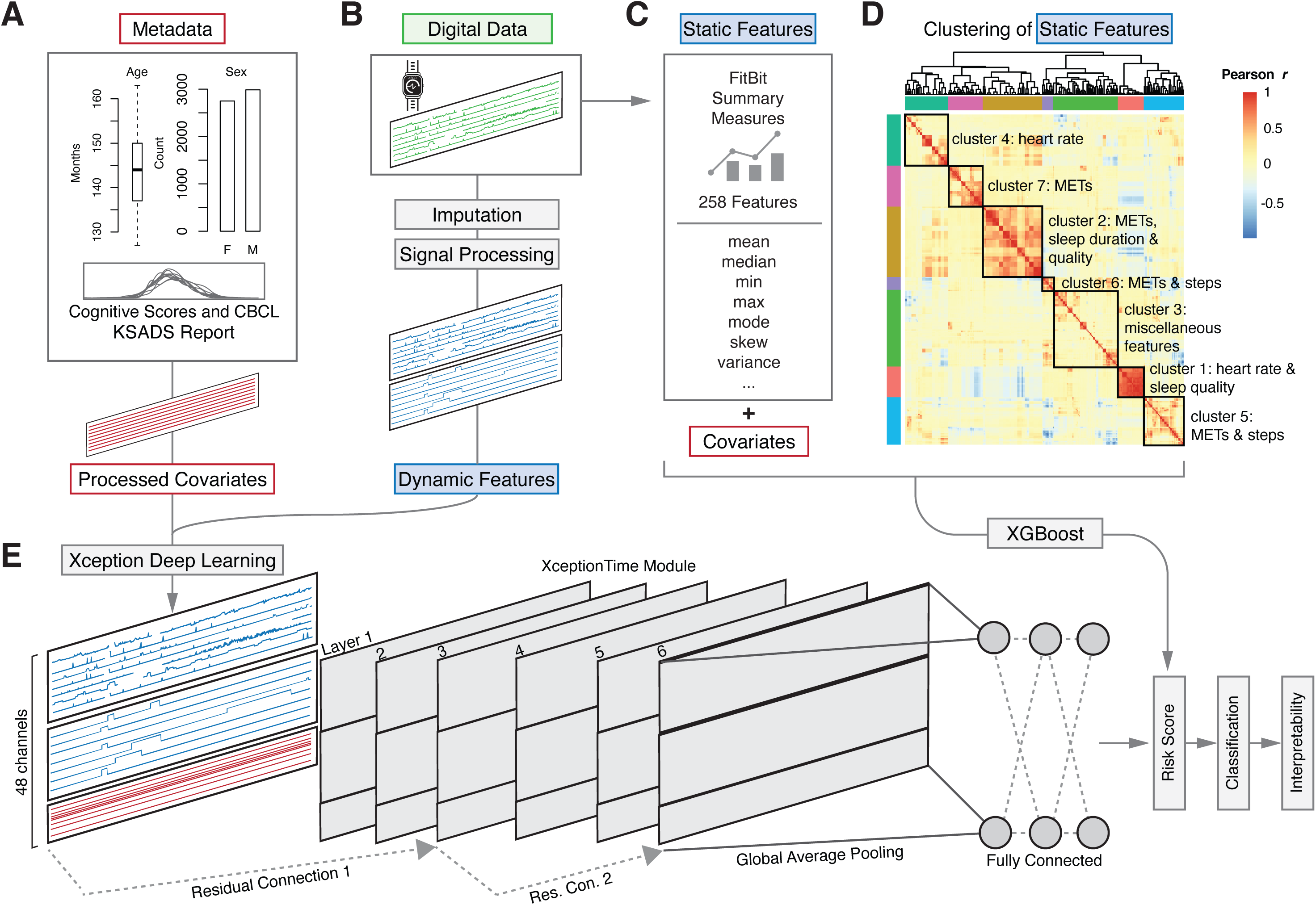
Workflow for data processing, feature engineering, and model architecture. **A)** ABCD cohort metadata including various demographic features, cognitive test scores, and clinical characteristics are used as covariates and represent the input features used in our baseline comparison model. Features shown in this plot correspond to the filtered set of individuals with wearable data. **B)** Digital data collected by wearable biosensors are used to generate dynamic features after signal processing and imputation steps. Together with the processed covariates, these time series features represent the input features for the dynamic model. **C)** Summary statistics applied to digital data collected by wearables are used to generate a total of 258 static features. In addition to the covariates, these are the input features used in the static model. The static model leverages the machine learning framework, XGBoost, for downstream tasks such as risk score generation, classification, model interpretability, and wearable GWAS. **D)** Hierarchical clustering of the static features yields seven distinct physiological clusters of wearable data. **E)** The dynamic model is based on the Xception deep learning framework, and uses the generated 48 channels from the dynamic features and covariates as input into a convolution-like model. The architecture consists of six inception layers and residual connections. Global average pooling and a fully connected layer allow for similar downstream tasks as mentioned in C).

### Generating intermediate phenotypes from wearable-derived data

We processed data obtained from FitBit smartwatches, which comprise measurements of heart rate, calories, activity intensity, steps, metabolic equivalents, sleep level and sleep intensity (**Fig. 1C**)^21^. These measurements quantify an individual’s physiological processes and their real-time changes in response to environmental stimuli, and can thus provide key information about an individual’s behavior.

To reconstruct the full spectrum of an individual’s behavioral functioning from these data, we applied two different feature engineering techniques, allowing us to generate wearable-derived dynamic and static features, which we consider as intermediate phenotypes. The dynamic features preserve the time-varying nature of the original data as a time series, enabling sequential and temporal patterns of the data to be retained. In contrast, the static features summarize patterns of the digital data to produce time-invariant, quantitative features that are commonly used in downstream modeling^14,22^.

To generate dynamic features, we performed signal imputation and processing after filtering the individuals with sparse data, and obtained 48 channels of time series (**Figure 2B**). Compared to the static features, this further processing allowed us to preserve both local and global temporal patterns potentially relevant to characterizing behavior and neurological response to stimuli^8,23^.

To generate static features, we first collected a total of 49 FitBit summary-based features. We next applied descriptive statistics (e.g. mean, median, etc.) to each of these features and generated a total of 258 static features for each individual (**Fig. 2C**)^14,24^. We then grouped these static features into seven main clusters, each of which summarizes different aspects of physiological and behavioral processes, such as heart rate, sleep duration and quality, metabolic intake or physical activity (**Fig. 2D**).

Altogether, static and dynamic features represent the physiological and behavioral profiles of the adolescents, and can be leveraged as intermediate phenotypes in a wide range of analyses to better characterize psychiatric phenotypes, such as generating disorder-specific probability risk scores, macrophenotype classification, model interpretability, and biomarker identification via wearable GWAS.

### Predicting psychiatric phenotypes from wearable-derived intermediate phenotypes

To demonstrate the validity of static and dynamic features as clinically relevant intermediate phenotypes and to evaluate their utility as a diagnostic tool, we employed these features in an array of classification tasks to identify individuals with either an externalizing (ADHD) or internalizing (anxiety) disorder from their typically developing peers. We selected ADHD and anxiety due to their high prevalence in adolescents, which is mirrored in the cohort (**Fig. 1B**)^25^.

We applied a gradient boosting machine learning algorithm, XGBoost, for classification tasks using static features (**Fig. 2C-D**)^26^. On the other hand, to fully leverage the time series nature of the dynamic features, we used a convolutional neural network for time series, featuring depthwise separable convolution, called Xception **(Fig. 2E)**^27^. Variable convolutional filters and residual (skip) connections, coupled with efficient parametrization, allow information encoded in both small and large receptive fields to be more optimally leveraged. Practically, this framework takes into account local and global patterns of physiology and behavior when performing downstream classification of psychiatric disorders. In both modeling approaches we included covariates that accounted for demographic features, family history of disorders, and other clinical information (**Fig. 2A**). To assess the benefit, in terms of model performance, of including wearable-derived data, we also trained a baseline model using just the covariates, which served as a comparison to the models including static or dynamic features. In practice, this comparison allowed us to determine whether wearable-derived features can improve diagnostic accuracy relative to that achievable using only a widely used broadband behavior rating scale.

After data filtering, we first used static features to classify 216 individuals with ADHD (an externalizing disorder) versus 1,737 of their typically developing peers (healthy controls) (**Fig. 3A)**. Using static features with XGBoost, we achieved an average area under the receiver operating characteristic curve (AUROC) of 0.87 and precision of 0.79. When using the dynamic features and Xception, we were able to achieve an average AUROC of 0.89 and precision of 0.83. The baseline model consisting of only the covariates achieved an average AUROC of 0.83, suggesting that the inclusion of wearable-derived features facilitates a clinically meaningful improvement in diagnostic accuracy. This improvement between the baseline and dynamic features model demonstrates statistical significance (one-sided t-test between baseline and dynamic features model, *p* value = 0.0022).

**Figure 3.**
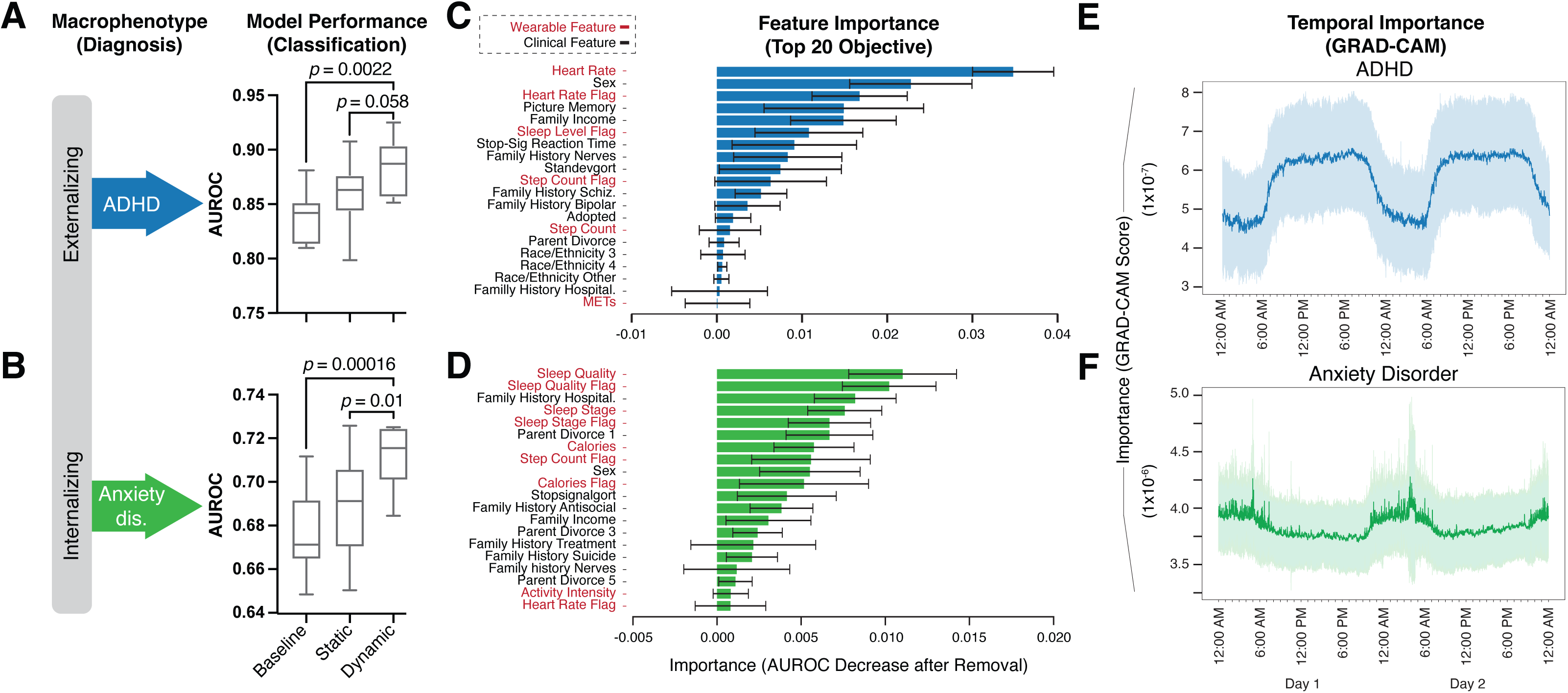
Performance and interpretability of psychiatric phenotype classification models. **A-B)** Model performance for baseline, static, and dynamic models employed for classifying individuals with ADHD (blue, top) or individuals with anxiety disorder (green, bottom) versus healthy controls. *P* values were calculated using one-sided t-test. **C-D)** Feature importance based on ablation studies for the dynamic model for ADHD (blue, top) and anxiety disorder (green, bottom) classification. Wearable-derived dynamic features are shown in red font and clinical features (covariates) are shown in black font. Feature importance is equivalent to the decrease in model performance (AUROC) after removal of the given feature. **E-F)** Temporal importance during a 48-hour period for dynamic features in ADHD (blue, top) or anxiety disorder (green, bottom) classification based on the GRAD-CAM interpretability module. Importance is represented as the GRAD-CAM score, based on each time points contribution towards model performance.

Second, we evaluated the performance of our model using static or dynamic features in the classification of 666 individuals diagnosed with anxiety disorder (internalizing disorder) versus 1,737 of their typically developing peers (healthy controls) (**Fig. 3B)**. Here, we again repeated the use of the same modeling framework, i.e., static features with XGBoost and dynamic features with Xception, and compared it to the baseline covariate model. We found that static and dynamic features achieve an average AUROC of 0.69 and 0.71 and precision of 0.64 and 0.68, respectively. In both models, the performance was greater than that of the baseline model (average AUROC of 0.67), with the dynamic features model showing the most significant performance improvement (one-sided t-test between baseline and dynamic feature model, *p* value = 0.00016). Overall, the fact that the models using dynamic features achieved the highest performance suggests the usefulness of the temporal patterns intrinsic to wearable-derived data towards understanding behavior.

### Interpreting wearable features prioritized by the deep learning model

Deep learning methods are typically characterized by complex internal structures that cannot be easily interpreted by humans. While maximizing the classification accuracy is one crucial aspect for characterizing complex phenotypes, understanding which features are most important in terms of their individual contribution to performance is also critical. To this end, we utilized ablation techniques to determine the relative contribution of each individual feature to model performance. For the ADHD classification task, heart rate was the most important feature (largest change in AUROC), followed by other dynamic features (i.e., sleep, steps, METs) as well as covariates such as demographics, family history, and cognitive scores from picture memory and stop-signal reaction time tests (**Fig. 3C**).

On the other hand, the ablation study for the anxiety classification task revealed a different set of important features. In this case, sleep quality and stage, calories, and step count were the most important dynamic features, whereas heart rate features, which were extremely important for classifying ADHD, were not prioritized in the anxiety model (**Fig. 3D**). Additionally, while the anxiety model prioritized some covariates that were relevant also for the ADHD model (e.g., sex, family history, and family divorce), cognitive scores from tests such as picture memory did not appear to be important for the identification of individuals diagnosed with anxiety, consistent with theory-driven accounts of neurocognitive aspects of anxiety disorders^28^.

Additionally, we assessed the importance for model performance of dynamic features at various times throughout the day by adapting gradient-weighted class activation mapping (Grad-CAM) strategies^29^. We calculated the relative importance of each time point in our dynamic features. For ADHD, we observed enriched significance of the heart rate dynamic feature around the early afternoon, potentially suggesting stronger behavioral differences between adolescents with ADHD and their typically developing peers (healthy controls) during this time of day (**Fig. 3E**). This is consistent with clinical research demonstrating time-of-day effects on ADHD symptom expression^30^. In contrast, sleep-related dynamic features during the night are much more informative in classifying anxiety, consistent with clinical expectations (**Fig. 3F**)^31^. Together, the ablation studies suggest a role for wearable-derived features to not only serve as quantitative intermediate phenotypes, but also to more closely reveal insights into the behavioral and physiological temporal patterns related to categorical macrophenotypes.

### Using wearable-derived features as intermediate phenotypes for wearable GWAS

Our accurate classification of both internalizing and externalizing psychiatric phenotypes based on wearable-derived features suggests that these features can serve as useful intermediate phenotypes and may be leveraged to identify genetic associations with psychiatric conditions (**Fig. 4**). To this end, we first focused specifically on ADHD, given the higher predictive power observed with our models (**Fig. 3A-B**) and its higher estimated heritability compared to anxiety (77-88% vs. 30-60%)^32,33^. We selected 1,191 individuals (137 individuals with ADHD and 1,054 healthy control individuals) with genetic and wearable data available, and performed a GWAS using the continuous prediction scores obtained from our wearable modeling framework. In practice, these scores represent risk probabilities for ADHD^34,35^. We identified 10 genome-wide (*p* value < 5·10^-8^) significant loci and 21 psychiatric or brain-related genes (**Fig. 4B**). Three of the identified genes (*ADORA3*, *PSMD11* and *DLG4*) have been previously associated with ADHD, bolstering the overall functional significance of the results^36–38^. Furthermore, several of these loci overlap with previously reported GWAS SNPs related to ADHD, neuroticism, sleep disruption and other clinically relevant traits (**Fig. 4B**). Note that here we used a continuous risk score as a univariate response variable for the GWAS. In comparison, when performing a traditional case‒control GWAS for ADHD on the same set of individuals using the binary diagnostic label (presence/absence of disorder) as response variable, we did not identify any significant loci (**Fig. 1A**, **Fig. 4A**). This result is consistent with the higher statistical power of continuous measurements over dichotomized (i.e., binary) traits, and with the findings that intermediate phenotypes show higher genetic penetrance compared to macrophenotypes^7,39–41^.

**Figure 4.**
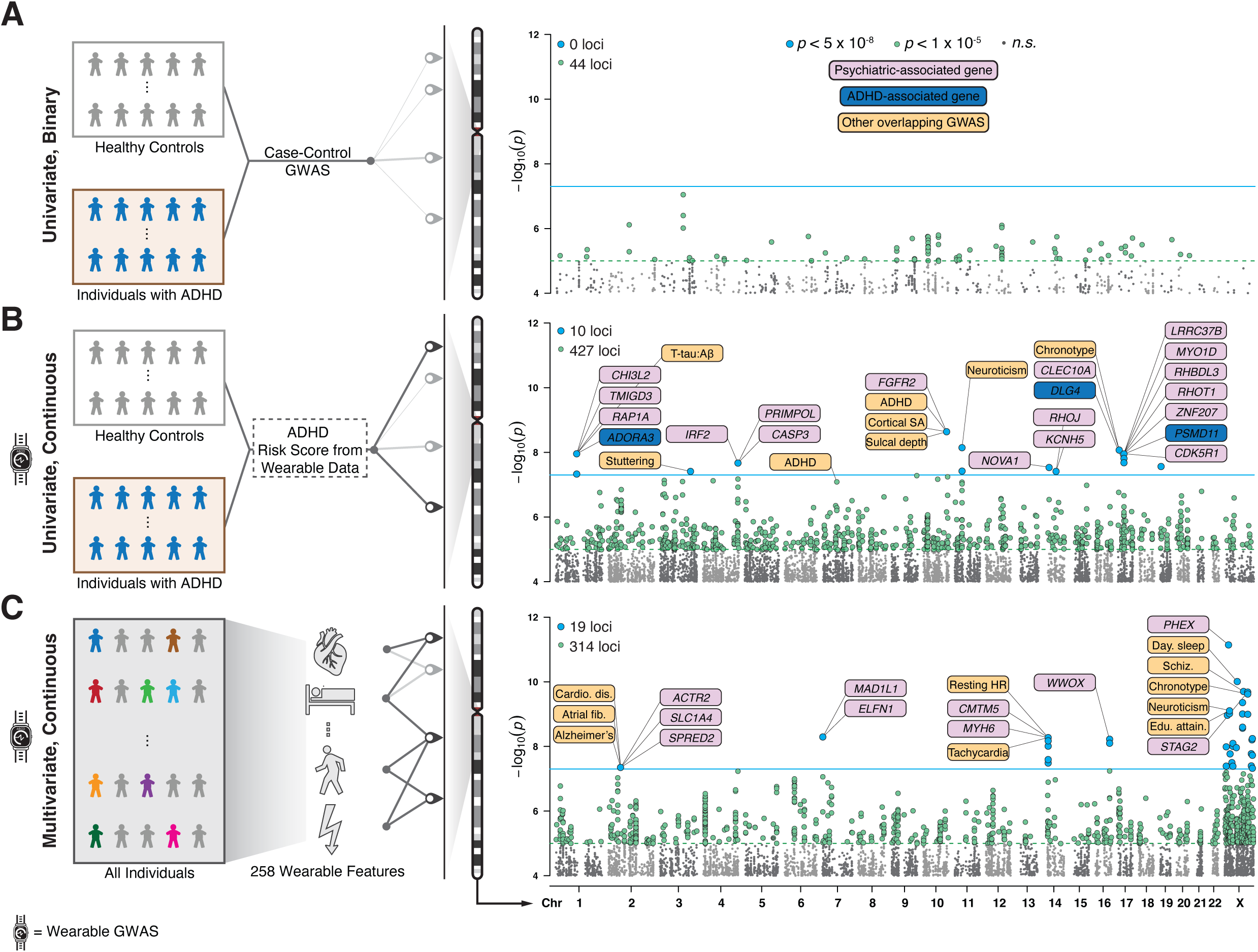
Manhattan plots summarizing the results of the univariate and multivariate wearable GWAS. **A)** Left panel: Case-control GWAS on 1,191 individuals from the ABCD cohort. We employed the clinical diagnosis label as the univariate binary response variable for the GWAS (1 = “individual with ADHD”, *n* = 137; 0 = “healthy control individual”, *n* = 1,054). Right panel: Manhattan plot showing the -log_10_ *p* value of association between the genetic variants and the univariate binary response variable. No genetic variants passed the genome-wide significance threshold (*p* value < 5·10^-8^; blue line). Genetic variants with a suggestive *p* value (< 10^-5^) are represented as green dots. **B)** Analogous representation to panel A) using the wearable-derived risk scores for ADHD as univariate continuous response variable. The GWAS was performed on the same set of 1,191 individuals and using the same set of covariates as in panel A). 10 and 427 loci passed the *p* value thresholds of 5·10^-8^ and 1·10^-5^, respectively. Loci chr1:111,372,165-111,482,359, chr17:7,101,607-7,101,608, and chr17:32,256,997-32,283,356 are proximal to genes *ADORA3* (72 Kb), *DLG4* (86 Kb) and *PSMD11* (174 Kb)(highlighted in dark blue) respectively, which have been previously associated with ADHD. Other proximal genes related to other psychiatric disorders are highlighted in pink (evidence obtained from OpenTargets). Brain-related traits associated with genetic variants overlapping the ten genome-wide significant loci are highlighted in orange. GWAS associations were obtained from the EBI-NHGRI GWAS catalog. **C)** Analogous representation to panel A) using the 258 wearable-derived static features as multivariate continuous response variable. The GWAS was performed on a set of 2,410 individuals (both healthy controls and individuals with any disorder). 19 and 314 loci passed the *p* value thresholds of 5·10^-8^ and 1·10^-5^, respectively. Neuropsychiatric-related genes proximal to the identified loci are shown in pink. Brain-, heart-, and sleep-related traits with associated variants overlapping the 19 loci are highlighted in orange.

While the analysis above collapses wearable-derived features into a single continuous variable that summarizes the risk score for a particular disorder (i.e., GWAS with a univariate response), it is also possible to directly use the full set of wearable-derived features as a more comprehensive and richer phenotype to represent the continuum of psychiatric disorders and their latent manifestations (i.e., GWAS with a multivariate response). In fact, these features can collectively capture behavioral patterns by measuring physiological processes and their real-time changes in response to environmental stimuli, and unlike disease risk scores, are not restricted to a specific cohort of individuals^42,43^. In what follows, we employed a multivariate nonparametric test of association to regress the vector of wearable-derived features on the genotype of each genetic variant, employing a larger cohort that spans healthy controls and individuals with any psychiatric disorder (*n* = 2,410)^44^. From this novel type of GWAS we identified 19 significant loci and 31 genes with a documented role in neurodevelopmental and psychiatric disorders (**Fig. 4C**). Many of these loci overlap previously identified GWAS SNPs related to heart, sleep, metabolism, and brain traits (**Fig. 4C**). This aligns with the close association between physiological functions, the central nervous system, and individual behavior.

### Functionally dissecting and interpreting novel wearable GWAS loci

To further investigate the loci identified by the behavioral GWAS, we dissected the variants using a battery of publicly available genomic resources^45,46^. Many of these loci overlap either GTEx expression quantitative trait loci (eQTLs) or ENCODE candidate cis-regulatory elements (cCREs), suggesting a link between the biochemical activity of these variants and their functional impact on macrophenotype. We also explored the impact of these loci beyond behavioral traits and their relationship with clinical psychopathology. For example, behavioral traits significantly associated with a specific genetic variant may correlate with clinical symptoms of a specific psychiatric cohort. Indeed, in some cases we show that the genetic variant in question is also differentially enriched between that specific psychiatric cohort and healthy individuals.

For instance, we found the minor allele (G) at rs365990 to be significantly associated with an increase in mean heart rate and a decrease in interday heart rate variation (**Fig. 5A, left**). The variant, missense for *MYH6*, had been previously linked to atrial fibrillation, ventricular tachycardia and resting heart rate, and the entire locus shows a significant enrichment of chromatin features in heart samples compared to other tissues and organs^47–50^. We also found the same allele to be enriched in the bipolar/psychotic disorder cohort compared to healthy controls (**Fig. 5A, right**). This cohort included youth meeting criteria for bipolar or unspecified psychotic spectrum disorder, and such severe pathology is known to be associated with characteristic irregularities in heart activity^51–53^. SNP rs365990 is also a GTEx eQTL for the *CMTM5* gene, which is highly expressed in brain subregions and has been implicated in stress response and childhood adversity, further supporting the relevance of this locus for psychiatric conditions in addition to heart pathophysiology^46,54^.

**Figure 5.**
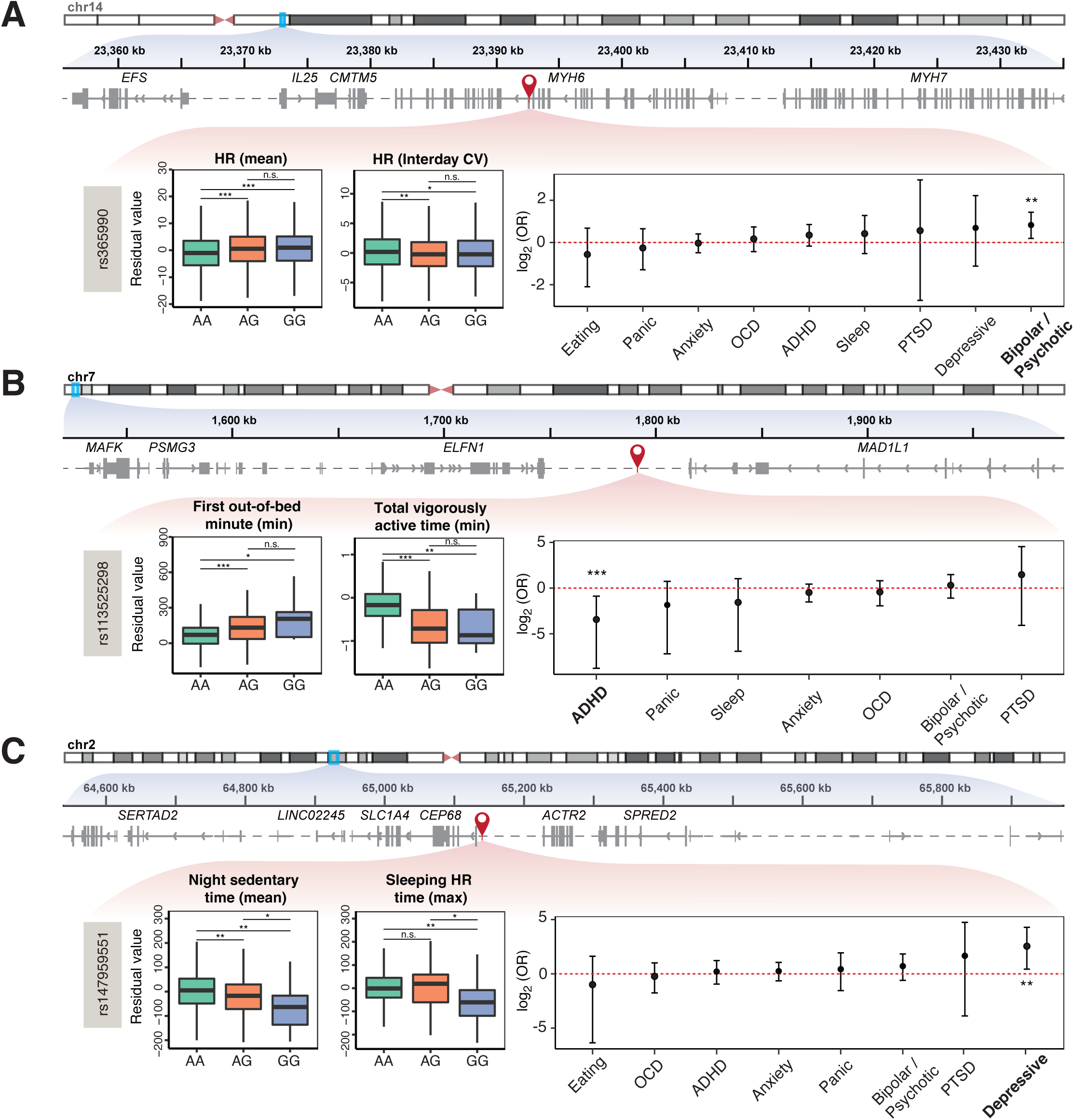
Exploring the genetic-physiological-psychiatric axis with wearable GWAS. **A)** Left panel: rs365990 (chr14:23,392,602, A/G) is located in exon 25 of *MYH6*, and is associated with changes in wearable-derived heart rate features (multivariate GWAS *p* value = 5.33E-09). The boxplots show distributions of covariate-adjusted mean and interday coefficient of variation (CV) for heart rate across genotype groups at rs365990 (AA *n* individuals = 1,228; AG *n* individuals = 1,509; GG *n* individuals = 519). *p* values (two-sided Wilcoxon Rank-Sum test) for each pairwise comparison are also displayed, encoded as follows: *** (*p* ≤ 0.001), ** (0.001 < *p* ≤ 0.01), * (0.01 < *p* ≤ 0.05), n.s. (*p* > 0.05). For visualization purposes, outliers are not shown. Right panel: enrichment, displayed as odds-ratio (log2 OR; y axis) of the minor allele (G) in individuals with different psychiatric disorders (x axis) compared to healthy controls. OR estimates and 95% confidence interval (error bar) are displayed. The red horizontal dashed line indicates no enrichment. The G allele is significantly more enriched in individuals with bipolar/psychotic disorder compared to healthy controls (two-sided Fisher test *p* value: 8.00E-03; FDR-adjusted *p* value: 7.00E-02). **B)** Similar representation for rs113525298 (chr7:1,791,353; AA *n* individuals = 2,294; AG *n* individuals = 101; GG *n* individuals = 15). rs113525298 is located 125 Kb from *ELFN1*, a gene that encodes a postsynaptic protein involved in the temporal dynamics of interneuron recruitment^65,66^. *Elfn1* mutant mice exhibit hyperactivity that is treatable by psychostimulant medication^55,56^. The G allele at rs113525298 is associated with increased minimum number of first-out-of-bed minutes and decreased minimum number of total-vigorously-active minutes (multivariate GWAS *p* value = 5.10E-09), and is significantly more enriched in healthy controls compared to individuals with ADHD (two-sided Fisher test *p* value: 9.00E-04; FDR-adjusted *p* value: 6.00E-03). **C)** Similar representation for rs147959551 (chr2:65,140,366; AA n individuals = 2,279; AG n individuals = 117; GG n individuals = 14), located near a cluster of genes relevant for several psychiatric disorders, such as *ACTR2* (schizophrenia; 87 Kb), *SLC1A4* (schizophrenia, bipolar disorder, major depressive disorder; 117 Kb) and *SPRED2* (schizophrenia, OCD; 170 Kb)^67–77^. The G allele of rs147959551 is associated with decreased mean number of sedentary-time-at-night minutes and decreased maximum number of sleep-based-on-heart-rate minutes (multivariate GWAS *p* value = 4.47E-08), and is significantly more enriched in individuals with depressive disorder compared to healthy controls (two-sided Fisher test *p* value: 9.74E-03; FDR-adjusted *p* value: 7.80E-02).

In a similar fashion, we explored variants rs113525298 and rs147959551. The minor allele at rs113525298 is associated with prolonged periods in bed and shorter vigorously active time during the day, and appears at a lower frequency in the ADHD cohort compared to healthy individuals (**Fig. 5B**). This suggests a potential protective role of the allele against hyperactivity disorders, further supported by the proximity of the SNP to *ELFN1*, previously implicated in the pathophysiology of ADHD^55,56^. On the other hand, we found the minor allele at rs147959551 associated with shorter sedentary time at night and a shorter period of time identified as sleep based on heart rate, two features suggestive of sleep disruption (**Fig. 5C)**. The same allele is also enriched in individuals with depression disorder, consistent with growing evidence implicating sleep impairment as a transdiagnostic feature of many forms of adolescent psychopathology (**Fig. 5C, right)**^57,58^.

Overall, these results highlight how wearable-derived features can be leveraged as intermediate phenotypes in GWAS and enable the identification of genetic variants relevant to clinical psychiatry with significant effects on exhibited behavior in adolescents.

## Discussion

Psychiatric disorders have been traditionally described with diagnostic categories based on retrospective self-report of symptom sets. However, current efforts in the field are increasingly leveraging novel technologies to transition from retrospective self-reporting and fixed symptom sets to more dimensional conceptualizations, aiming to capture the complex and heterogeneous nature of psychiatric disorders for more accurate research into their underlying structure^6^. One approach to enhancing dimensional models is the use of intermediate phenotypes—quantitative traits linked more closely to a disorder’s underlying molecular pathways. Although intermediate phenotypes have been derived from cellular, tissue, and organ levels of information, computational strategies that generate useful intermediate phenotypes in the behavioral domain are currently limited. Wearable biosensors such as smartwatches offer a unique opportunity to objectively study psychiatric disorders in a non-invasive way by measuring their underlying physiological foundations of behavior over time.

Towards this end, we used wearable data to generate static and dynamic features that were employed by our AI modeling framework as intermediate phenotypes to distinguish between adolescents with and without psychiatric disorders. Models utilizing these wearable-derived intermediate phenotypes performed comparably to those based on more expensive data sources such as fMRI measurements^18,59^. To gain critical theoretical insights and inform treatment development efforts, we augmented the modeling framework with interpretability modules, allowing us to pinpoint temporal and functional regions of the time series that were highly correlated with overall disease state^60^. These interpretability modules have the potential to facilitate mechanistic studies that offer deeper insight into the underlying complexities of these disorders. For example, our interpretability modules revealed that heart rate time series held high importance in predicting ADHD. This finding aligns with the clinical manifestation of ADHD – affected children are characterized by episodes of heightened arousal that are often incongruent with environmental demands^61^. Conversely, the interpretability modules identified sleep intensity and quality as key predictors in our anxiety disorder models, in line with known disruptions in sleep patterns and circadian rhythms commonly seen in youth with anxiety disorders^62^.

Wearable-derived intermediate phenotypes are not just effective for detecting the presence of psychiatric disorders in individuals; they also serve as a valuable research tool for understanding the correspondence between behavior patterns and molecular attributes. This comprehensive approach helps to uncover the foundational elements of pathological behavior patterns. In this context, we focused on establishing links with genetics. Specifically, we showed that these intermediate phenotypes can serve as response variables in GWAS models. Their continuous nature enhances statistical power compared to categorical diagnostic labels. Furthermore, we took advantage of the features’ correlated structure to create multivariate response variables for GWAS. This strategy is statistically advantageous because it mitigates the multiple testing burden associated with evaluating the numerous (>250) independent features. Conversely, from a biological standpoint, these wearable GWAS allowed us to explore triaxial associations encompassing genetic, physiological, and psychiatric factors. Utilizing our framework, we successfully identified a significant association between a missense variant of the *MYH6* gene, which encodes the cardiac muscle myosin, and heart rate patterns. Heart activity receives complex inputs from the CNS, which implies behavioral influence and, in combination with our GWAS, supports the notion of a gene-behavior-disorder pathway^63^. Building on this finding, we discovered enrichment of the same genetic variant among individuals with bipolar/psychotic disorders, psychiatric conditions known to be associated with characteristic irregularities in heart activity^51^. While additional research is needed to confirm such associations, our findings resonate with the objectives of the RDoC initiative^6^. Specifically, wearable-derived intermediate phenotypes serve as objective markers of behavior, bridging lower-level biological systems like genetics to broader psychiatric disorders.

While we have employed these wearable-derived intermediate phenotypes in a targeted research context (i.e., to enhance a psychiatric GWAS), their broad applicability make them promising for other domains of health research. For example, the risk scores generated by our AI-modeling framework could be used to assess disorder severity, and the genetic variants identified by our wearable GWAS could be employed to construct more comprehensive polygenic risk scores for behavioral and psychiatric disorders. Unlike other diseases (e.g., cancer) where objective biomarkers are more common, psychiatry faces a significant barrier in treatment due to the lack of objective and sensitive screening methods^64^. Therefore, these physiological and genetic features could be leveraged as objective biomarkers to more accurately subtype patients within diagnostic categories, which in turn could help move towards precision treatment delivery in psychiatry.

Although the results presented in this study require further experimental validation, they illuminate the transformative potential of wearable devices combined with AI modeling frameworks for deepening our understanding of complex behavioral and psychiatric traits. We anticipate that further development of our AI modeling framework, coupled with an expanded array of wearable devices, could fundamentally transform how psychiatric disorders are measured and understood in both research and clinical settings. This could lead to more nuanced digital intermediate phenotypes and open new avenues for the study of human behavior.

## Data Availability

Data used in the preparation of this article were obtained from the Adolescent Brain Cognitive DevelopmentSM (ABCD) Study (https://abcdstudy.org), held in the NIMH Data Archive (NDA). This is a multisite, longitudinal study designed to recruit more than 10,000 children age 9-10 and follow them over 10 years into early adulthood. The ABCD Study is supported by the National Institutes of Health and additional federal partners under award numbers U01DA041048, U01DA050989, U01DA051016, U01DA041022, U01DA051018, U01DA051037, U01DA050987, U01DA041174, U01DA041106, U01DA041117, U01DA041028, U01DA041134, U01DA050988, U01DA051039, U01DA041156, U01DA041025, U01DA041120, U01DA051038, U01DA041148, U01DA041093, U01DA041089, U24DA041123, U24DA041147. A full list of supporters is available at https://abcdstudy.org/federal-partners.html. A listing of participating sites and a complete listing of the study investigators can be found at https://abcdstudy.org/consortium_members/. ABCD consortium investigators designed and implemented the study and/or provided data but did not necessarily participate in the analysis or writing of this report. This manuscript reflects the views of the authors and may not reflect the opinions or views of the NIH or ABCD consortium investigators.

https://nda.nih.gov/abcd

## Acknowledgements

Data used in the preparation of this article were obtained from the Adolescent Brain Cognitive DevelopmentSM (ABCD) Study (https://abcdstudy.org), held in the NIMH Data Archive (NDA). This is a multisite, longitudinal study designed to recruit more than 10,000 children age 9-10 and follow them over 10 years into early adulthood. The ABCD Study® is supported by the National Institutes of Health and additional federal partners under award numbers U01DA041048, U01DA050989, U01DA051016, U01DA041022, U01DA051018, U01DA051037, U01DA050987, U01DA041174, U01DA041106, U01DA041117, U01DA041028, U01DA041134, U01DA050988, U01DA051039, U01DA041156, U01DA041025, U01DA041120, U01DA051038, U01DA041148, U01DA041093, U01DA041089, U24DA041123, U24DA041147. A full list of supporters is available at https://abcdstudy.org/federal-partners.html. A listing of participating sites and a complete listing of the study investigators can be found at https://abcdstudy.org/consortium_members/. ABCD consortium investigators designed and implemented the study and/or provided data but did not necessarily participate in the analysis or writing of this report. This manuscript reflects the views of the authors and may not reflect the opinions or views of the NIH or ABCD consortium investigators.

